# The youth of tamale metropolis: understanding energy drink consumption, perceptions and related factors

**DOI:** 10.1101/2023.07.20.23292872

**Authors:** Williams Kobik, Paul Armah Aryee

## Abstract

Energy drinks have become a popular choice for young people seeking physical and cognitive boosts, with ingredients such as caffeine, taurine, and B vitamins aimed at improving academic, athletic, and alertness levels. However, the popularity of these drinks is also driven by low prices, taste, brand loyalty, and gendered marketing, with boys being more likely to consume them. Despite the supposed benefits, energy drinks have been associated with high-risk behaviours, deaths, and adverse health effects, especially related to cardiovascular risk. Meanwhile, in Ghana, the use of energy drinks is on the rise. Hence, this study aimed to examine the prevalence and consumption pattern, perception, and factors associated with ED consumption among the youth of the Tamale Metropolis. The study was cross-sectional consisting of 541 participants. The group consisted of 340 males and 201 females, between the ages of 15 and 45. A questionnaire was utilized to obtain data on the respondents’ consumption patterns and perceptions of EDs, as well as their socio-demographic characteristics. The results of the study indicated that a large percentage of the respondents, 98.7%, had consumed energy drinks before, while 78.7% currently consume them. Respondents believed that energy drinks provided additional energy (81.00%) and reduced stress (62.30%). However, they also perceived side effects such as insomnia (60.60%) and restlessness (51.40%). Although the majority of respondents (83.4%) were unaware of the classification of energy drinks and their ingredients, side effects, and benefits. Age, marital status, level of education, work intensity, EDs served at gatherings, and knowledge of EDs was significantly associated with ED consumption (p < .05). Consumption was higher among those aged 26 to 35 years, singles, individuals with no formal education, and those with high work intensity. Energy drinks were consumed by the majority of the youth. The high consumption was also associated with low knowledge levels. It is recommended that public health and nutrition professionals should engage in further advocacy efforts to improve the youth’s perception of EDs in a positive manner. In addition, lawmakers should use legislation to influence consumption rates and safeguard the health of consumers.

## Introduction

Throughout history, humans have sought physical and cognitive enhancements to tackle challenging tasks. The rise of energy drinks (EDs) and their marketing strategies, aimed at boosting physical and cognitive performance, has led to a significant increase in consumption among young people [1]. Ingredients commonly found in energy drinks include caffeine, guarana, ginseng, taurine, glucose, L-carnitine, glucuronolactone, and B vitamins [2]. Despite their perceived benefits, factors such as low cost, taste, brand loyalty, and gendered marketing have contributed to their popularity. However, marketing campaigns geared towards masculinity have resulted in fewer girls consuming energy drinks than boys [3,4].

The energy drink market is one of the fastest-growing segments of the non-alcoholic beverage industry. Despite the notoriety of energy drinks being high in sugar and caffeine, their dangers have been largely overlooked. Although the European Union has stated that there is no evidence to support the argument that energy drinks pose a degree of toxicity to the extent of adverse health outcomes. However, case reports from around the world suggest otherwise [5–7].

Energy drinks have not only been associated with negative health effects and death but have also been linked to high-risk substance use and social deviance such as vandalism and reckless driving, resulting in accidents [8–11]. Acute effects include sudden cardiac death, high blood pressure, and endothelial dysfunction, which affect young people and those with underlying cardiovascular conditions. The usual ingredients found in energy drinks, including guarana, ginseng, taurine, and caffeine, have been associated with a range of adverse health outcomes [12]. Excessive sugar intake, which is often present in energy drinks, is also a contributor to overnutrition.

Sub-Saharan Africa is experiencing an upward trend in diet-related diseases, including hypertension, obesity, stroke, and heart disease, contributing to the rapid increase in the epidemic of non-communicable diseases. In Ghana, where energy drink consumption is on the rise, a survey revealed that 62% of student-athletes consume at least one can of energy drink per week. While the sale and use of energy drinks are not regulated in Ghana, developed countries such as France, Denmark, and Norway have banned energy drinks with high levels of caffeine and taurine. Some countries require that energy drinks are sold as medicinal products or with warning labels [13,14].

The overall aim of this study was to evaluate the youth’s perceptions, consumption habits, and factors linked with energy drink consumption in the Tamale Metropolis. This study will make clear enable the understanding of the consumption habits of energy drinks among youth in the Tamale Metropolis will contribute to raising awareness, prompt more research in the area and influence policies and public health actions.

## Methods

### Study design

The study design was a descriptive cross-sectional.

### Study area

The Tamale Metropolis is an urban area in the Northern Region with a population of about 360,579, 80.8% of which lives in urban areas and a size of 289.58 square miles [15]. Energy drinks are readily available through convenience shops and street vendors, and the area has a fast-growing business landscape where the trading of energy drinks thrives. Below is a map of the metropolis extracted from Google Maps.

### Study site

The data were collected in the central business district of the metropolis.

### Study population

The focus of this research was on individuals aged 15 to 45 years old. This age bracket was chosen for two reasons: Firstly, approximately 63.3% of the population aged 15 and above in the city are engaged in economic activities, making them financially capable of buying energy drinks [15]. Secondly, this group holds a particular belief about the advantages of these drinks, which impacts their decision to purchase them.

### Inclusion and Exclusion Criteria

The participants for this study were required to fall within the age range of 15 to 45 years and also be permanent residents of the Tamale metropolis. To verify residency, respondents were asked if they had lived in the metropolis for a minimum of five years. These criteria were set to ensure that the data collected was representative of the local population within this age bracket and who have had adequate exposure to the city’s environment and social dynamics.

### Sample size determination

The total population of youth (n) for this study, 383 was determined using Cochran’s formula. The confidence interval was set at 95% with the z-score (t) being 1.96. The margin of error (m) was 0.05 and the prevalence of energy drink consumption (p) was estimated to be 46.7% [16].

### Sampling technique

The Tamale Metropolis was purposively chosen as the study location, and respondents were selected for convenience. Specifically, study participants were chosen based on their presence in the central business district of the metropolis. Although convenience sampling was used to select participants, the process depended on the established inclusion and exclusion criteria.

### Data Collection and Quality Management

The study collected data on individual respondents in the Tamale metropolis through semi-structured interviews using a pretested-structured questionnaire administered via face-to-face interviews with the aid of a Computer Assisted Personal Interview (CAPI) device. The questionnaire covered demographic information, anthropometry, and perception, with the latter section probing respondents’ knowledge of energy drinks. The data was collected by enumerators who received a one-day training on questionnaire administration and translation into the local dialect. The questionnaire was divided into four sections covering sociodemographic characteristics, anthropometry, perception, and knowledge. Anthropometry involved measuring respondents’ height and weight, while perception sought to probe respondents’ understanding of energy drinks, including their ingredients, brands, benefits, and adverse effects. Based on the responses, a score was generated, and respondents’ knowledge was categorized as “poor,” “good,” or “excellent.”

### Data analysis

The collected data was transferred from the Kobo toolbox on mobile phones to Excel for cleaning and then to IBM Statistical Package for Social Sciences (SPSS version 21) for analysis. The consumption patterns of energy drinks were analyzed using frequencies and percentages. For continuous data, correlations, means, and standard deviations were used. The relationship between exposure variables such as demographics and perceptions and consumption patterns was examined using Chi-square tests. A p-value less than 0.05 was considered statistically significant for all analyses.

### Ethical considerations

The study followed ethical guidelines and obtained ethical approval from the Committee on Human Research and Publication Ethics (CHRPE) with Ref No: CHRPE/AP/256/22. Prior to data collection, informed consent was obtained from all participants, emphasizing their voluntary participation and the confidentiality of their information. Participants were provided with clear explanations regarding the purpose, procedures, and potential risks and benefits associated with the study. Participants were also assured of their right to withdraw from the study at any point without repercussions. To uphold privacy and confidentiality, personal identifiers were removed from the collected data, and data storage followed secure protocols. Only authorized researchers had access to the data, maintaining strict confidentiality throughout the study. The data was collected from 1^st^ to 31^st^ July 2022.

## Results

The data gathered explored respondents’ socio-demographic characteristics, knowledge, attitude, and practice regarding energy drinks. Analysis was done using various statistical tests, including T-tests, chi-square tests, Pearson’s r correlation, and logistic regression, with a significance level of 0.05.

### Socio-demographic characteristics of youth

The respondents had a mean age of 25.55 (*SD* = 8.11) years, with the majority (59.7%) falling in the 15-25 age range. Most respondents were male (62.8%), single (70.6%), practicing Islam (66.0%), and from the Dagomba ethnic group (55.1%). Around 44% of respondents were students, and a significant proportion had formal education. The majority (65.2%) engaged in labor-intensive work. An independent samples T-test revealed significant age differences between males and females (*t* (366.66) = -2.04, *p* < .001), with females (*M* = 36.51, *SD* = 8.92) being older than males (*M* = 24.98, *SD* = 7.55). However, there were no significant BMI differences between genders (*t* (539) = 1.623, *p* = .105). The chi-square test examined associations between gender and general characteristics. Marital status did not differ significantly by gender (*X*^2^(4, N = 541) = 4.67, *p* > 0.05). However, there were significant gender differences in the frequency of energy drink (ED) consumption (*X*^2^(1, N = 525) = 7.36, *p* = .007) and mixing EDs with other substances (*X*^2^(1, N = 413) = 10.65, *p* = .001). Employment status showed no significant gender differences (*X*^2^(3, N = 541) = 5.11, *p* = .164), but work intensity had a significant relationship with gender (*X*^2^(1, N = 541) = 8.01, *p* = .005). The level of education did not differ significantly between males and females (*X*^2^(4, N = 541) = 2.21, *p* = .697). Alcohol consumption was significantly higher among males than females (*X*^2^(1, N = 541) = 36.94, *p* < .001), and there were no female smokers (*X*^2^(1, N = 541) = 14.83, *p* < .001) (Table 1).

**Table 1.** Socio-Demographic Characteristics of Sampled Youth of Tamale.

| Variable | Total (541) | Male (340) | Female (201) | P-value |
| --- | --- | --- | --- | --- |
| **Age (yrs.), *n*(%)** |  |  |  | .021 |
| 15-25 | 323 (59.70) | 213 (39.40) | 110 (20.30) |  |
| 26-35 | 128 (23.70) | 82 (15.20) | 46 (8.50) |  |
| 36-45 | 90 (16.60) | 45 (8.30) | 45 (8.30) |  |
| **Age, mean (SD)** | 25.55 (8.11) | 24.98 (7.55) | 36.51 (8.92) | .034 |
| **Weight, mean (SD)** | 61.65 (9.08) | 63.69 (8.31) | 58.3532 (9.39) | <.001 |
| **Height, mean (SD)** | 1.64 (.10) | 1.66 (0.11) | 1.6019 (.07) | <.001 |
| **BMI, mean (SD)** | 23.19 (3.86) | 23.40 (3.85) | 22.8418 (3.86) | .105 |
| **Marital status, *n*(%)** |  |  |  | .335 |
| Never Married | 382 (70.61) | 249 (73.20) | 133 (66.20) |  |
| Married/Cohabiting | 141 (26.06) | 82 (24.10) | 59 (29.40) |  |
| Separated | 8 (1.48) | 5 (1.50) | 3 (1.50) |  |
| Divorced | 2 (.37) | 1 (.30) | 1 (.50) |  |
| Widowed | 8 (1.48) | 3 (.90) | 5 (2.50) |  |
| **Ethnicity, *n*(%)** |  |  |  | .001 |
| Dagomba | 298 (55.10) | 211 (39.0) | 87 (16.10) |  |
| Others | 243 (44.90) | 129 (23.8) | 114 (21.10) |  |
| **Religion, *n*(%)** |  |  |  | .059 |
| Christianity | 181 (33.50) | 100 (18.50) | 81 (15.00) |  |
| Islam | 357 (66.00) | 238 (44.00) | 119 (22.00) |  |
| Traditional | 3 (.50) | 2 (.40) | 1 (.10) |  |
| **Employment status, *n*(%)** |  |  |  | .164 |
| Student | 228 (42.10) | 134 (39.40) | 94 (46.80) |  |
| Unemployed | 49 (9.10) | 28 (8.20) | 21 (10.40) |  |
| Self-employed | 143 (26.40) | 99 (29.10) | 44 (21.90) |  |
| Employed | 121 (22.40) | 79 (23.20) | 42 (20.90) |  |
| **Work intensity, *n*(%)** |  |  |  | .005 |
| Yes | 353 (65.20) | 237 (69.70) | 116 (57.50) |  |
| No | 188 (34.80) | 103 (30.30) | 85 (42.30) |  |
| **Education level, *n*(%)** |  |  |  | .697 |
| None | 44 (8.10) | 29 (8.50) | 15 (7.50) |  |
| Primary | 28 (5.20) | 19 (5.60) | 9 (4.50) |  |
| JHS | 80 (14.80) | 49 (14.40) | 31 (15.40) |  |
| Secondary/vocational | 184 (34.00) | 121 (35.60) | 63 (31.30) |  |
| Tertiary | 205 (37.90) | 122 (35.90) | 83 (41.30) |  |
| **Alcohol intake, *n*(%)** |  |  |  | <.001 |
| Yes | 90 (16.60) | 82 (24.10) | 8 (4.00) |  |
| No | 451 (83.40) | 258 (75.90) | 193 (96.00) |  |
| **Smoking, *n*(%)** |  |  |  | <.001 |
| Yes | 24 (4.40) | 24 (4.40) | 0 (.00) |  |
| No | 517 (95.60) | 316 (92.90) | 201 (100.00) |  |

### The prevalence of energy drink consumption

Figure 1 illustrates that 98.7% of youth in Tamale have ever consumed an energy drink, and 78.7% of them are current consumers (Fig 1).

**Fig 1.**
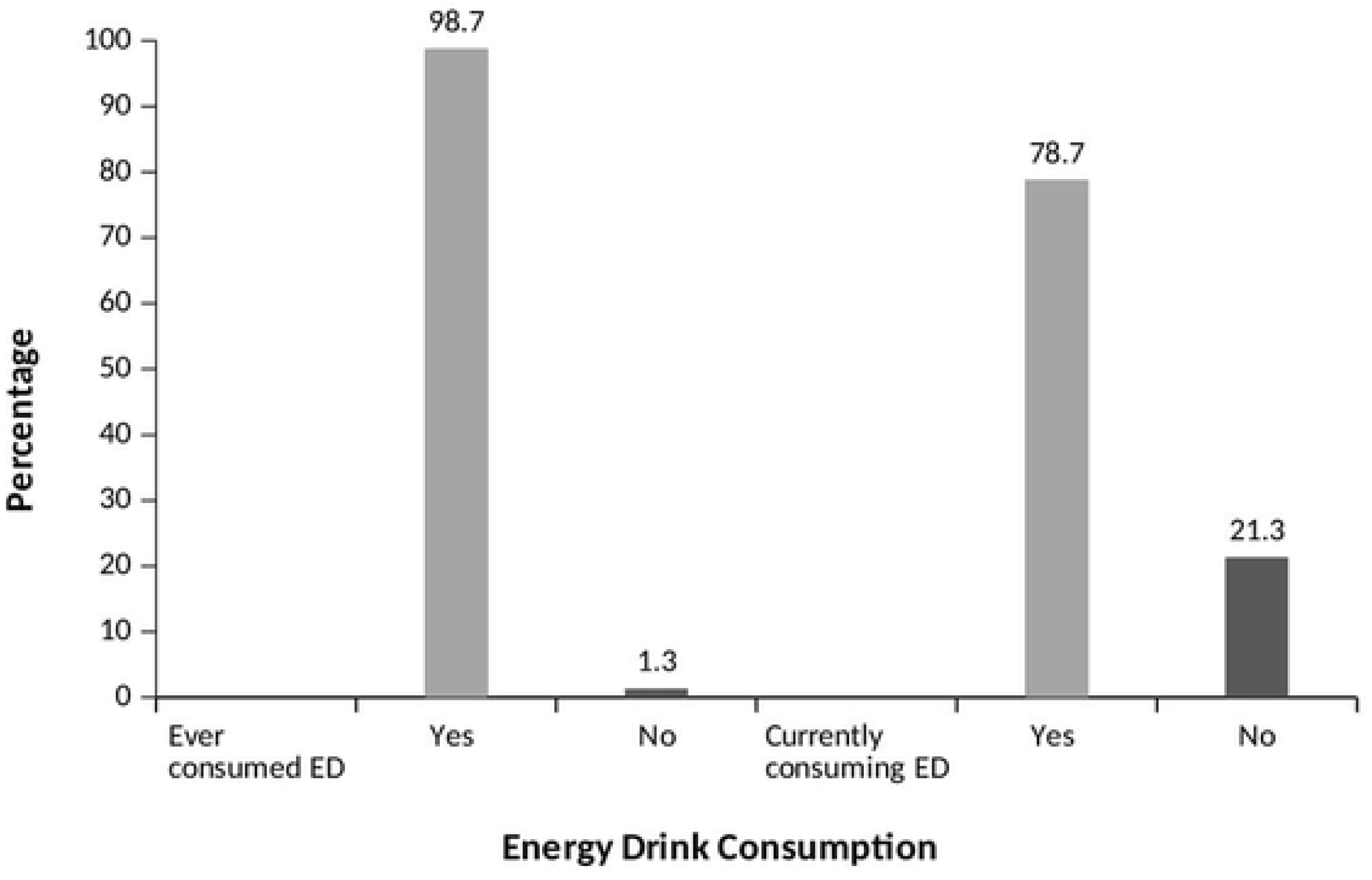
Prevalence of Energy Drink Consumption.

### Consumption pattern of energy drinks

The analysis of the data revealed that the highest frequency of energy drink consumption among the study sample was weekly (70.9%), with smaller proportions consuming daily (36.9%) or monthly (2.2%) (Table 2). The majority of consumers (93.9%) consumed one bottle per sitting, while smaller proportions consumed two (4.1%), three (1.2%), or four (0.7%) bottles/cans. The evening was the preferred time for consumption (55.0%), and most consumers (62.0%) had been consuming energy drinks for 2 to 3 years without a specific reason (Table 2). Fig 2 shows that the primary reasons for consuming energy drinks were to reduce stress and tiredness (60.3%), followed by seeking an energy boost (34.1%) and the desire to stay awake (29.5%). Smaller proportions mentioned other reasons such as increasing concentration, boosting appetite, refreshment, pain relief, sexual endurance, aiding digestion, seeking a high, enjoying the taste, and facilitating weight loss. Only 31.26% of respondents encountered energy drinks at social gatherings. The majority encountered them at weddings (50.3%), while 39.4% encountered them at outdoor events, and a smaller proportion encountered them at parties (Fig 3 and 4). Among the respondents, 25.9% experienced side effects. Insomnia was the most commonly reported side effect (55.1%), followed by post-consumption fatigue (18.1%), palpitations (16.8%), and restlessness (13.1%) (Fig 5 and 6).

**Fig 2.**
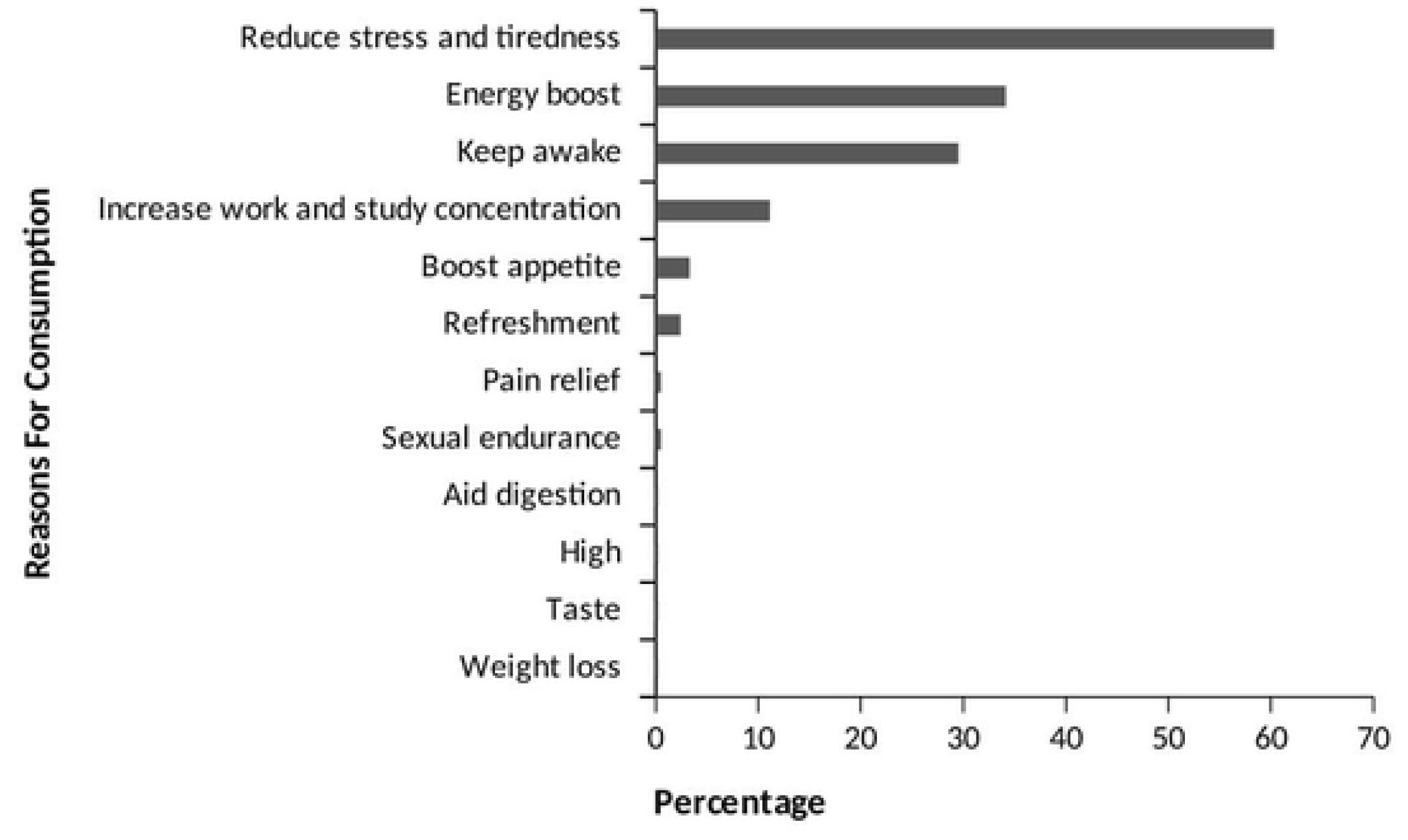
Reasons for Consuming Energy Drinks.

**Fig 3.**
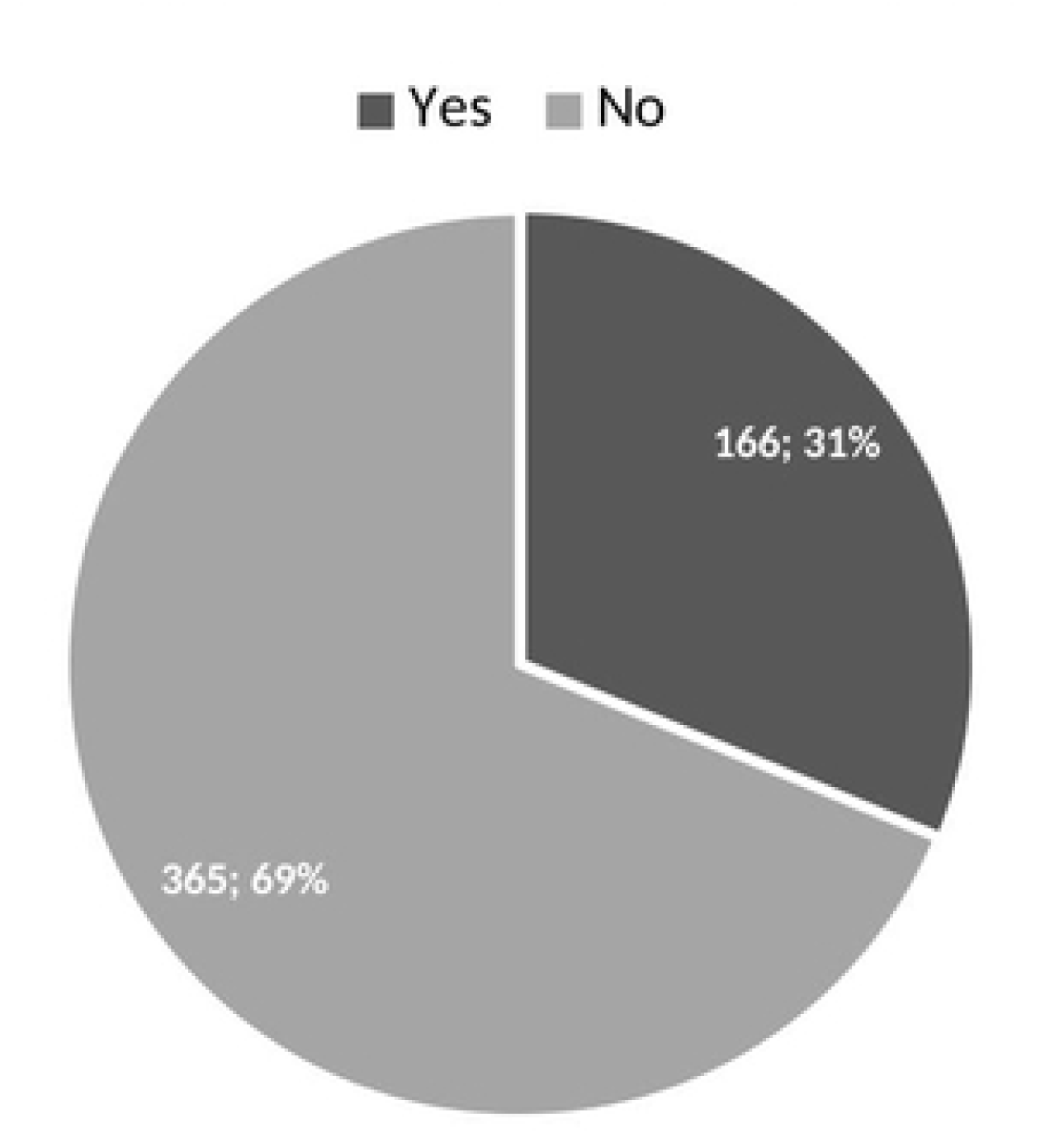
The Proportion of Energy Drink Sightings at Social Gatherings.

**Fig 4.**
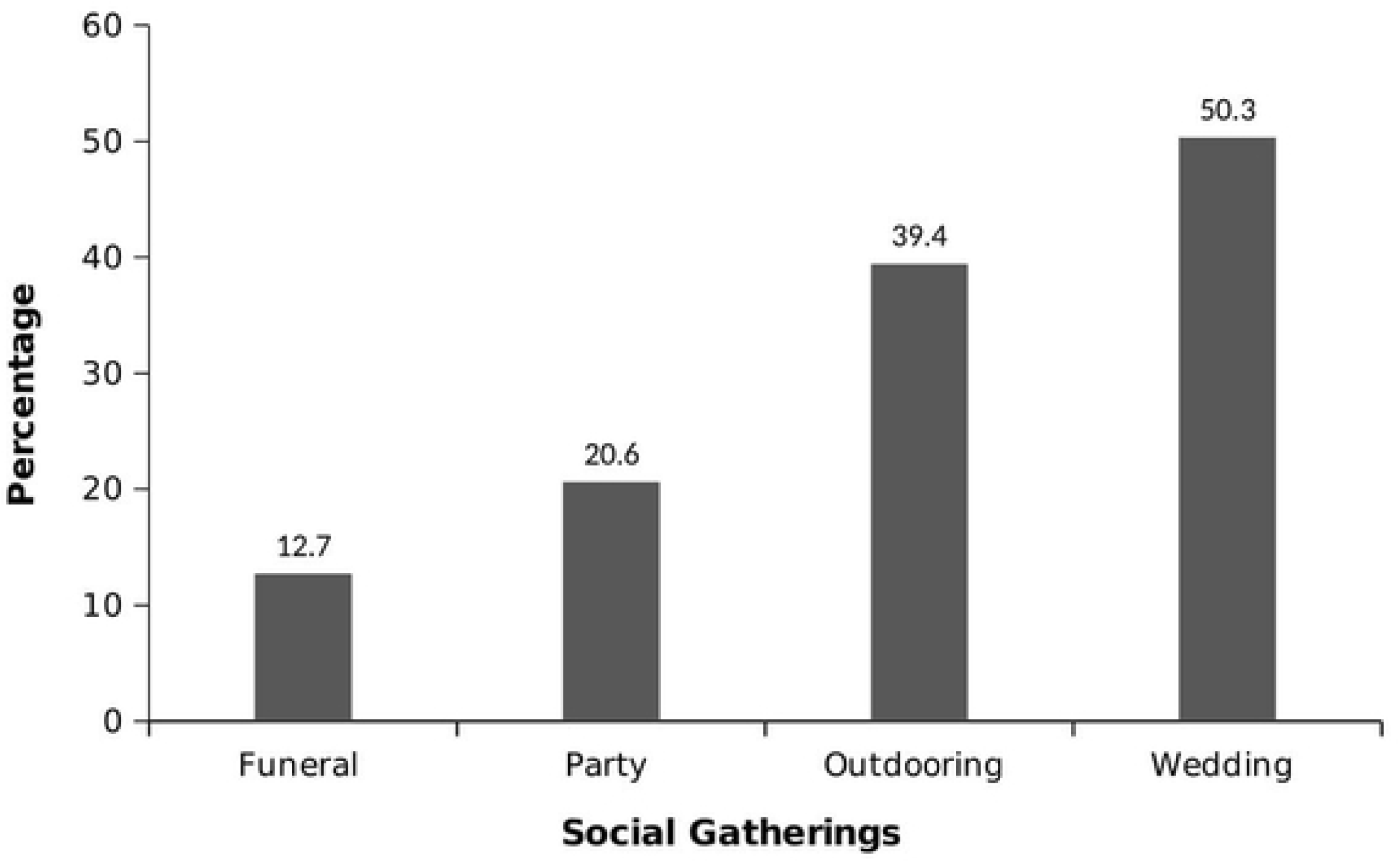
Social Gatherings Where Energy Drinks Have Been Sighted.

**Fig 5.**
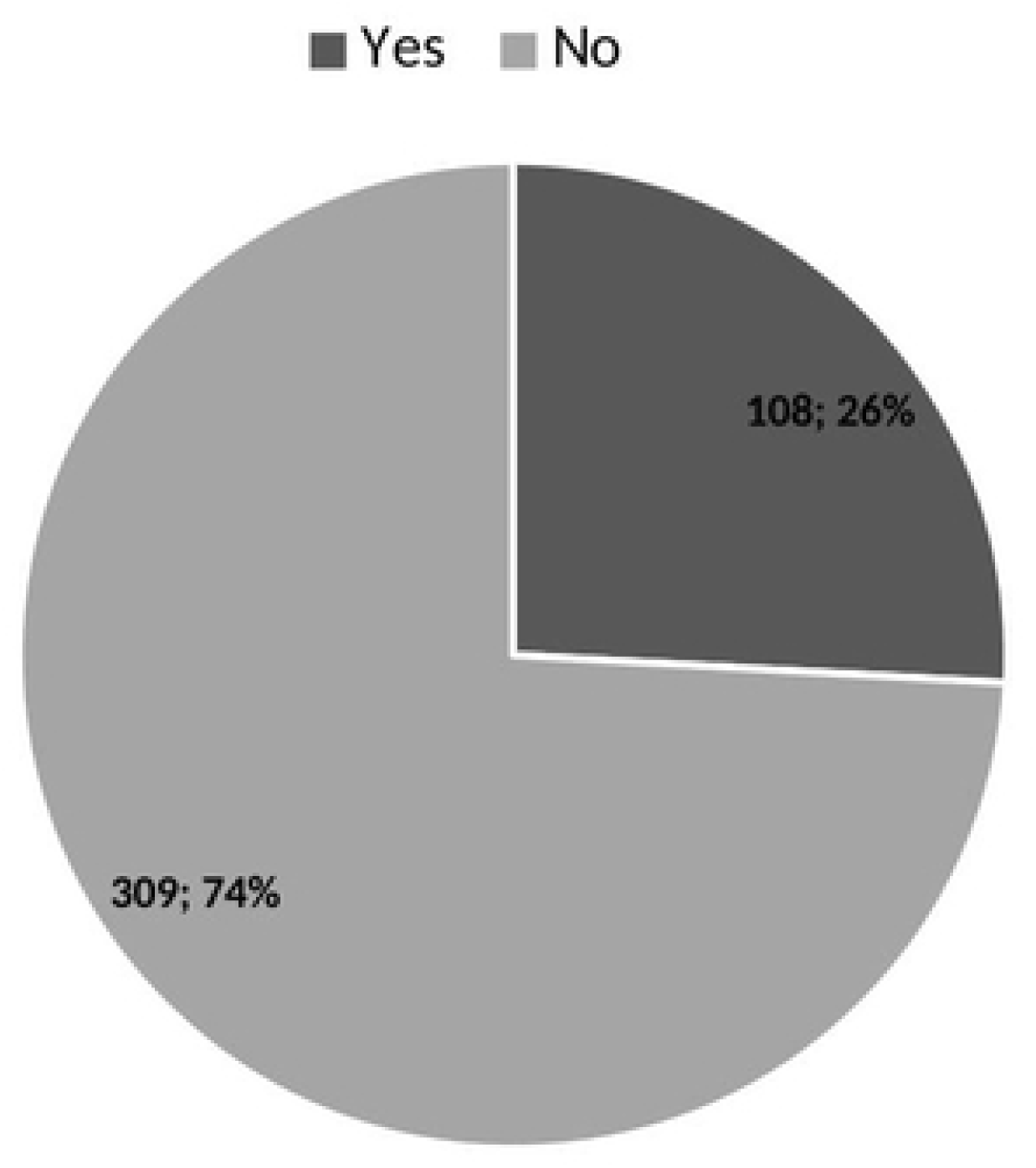
The Proportion of Energy Drink Users Who Have Experienced Side Effects.

**Fig 6.**
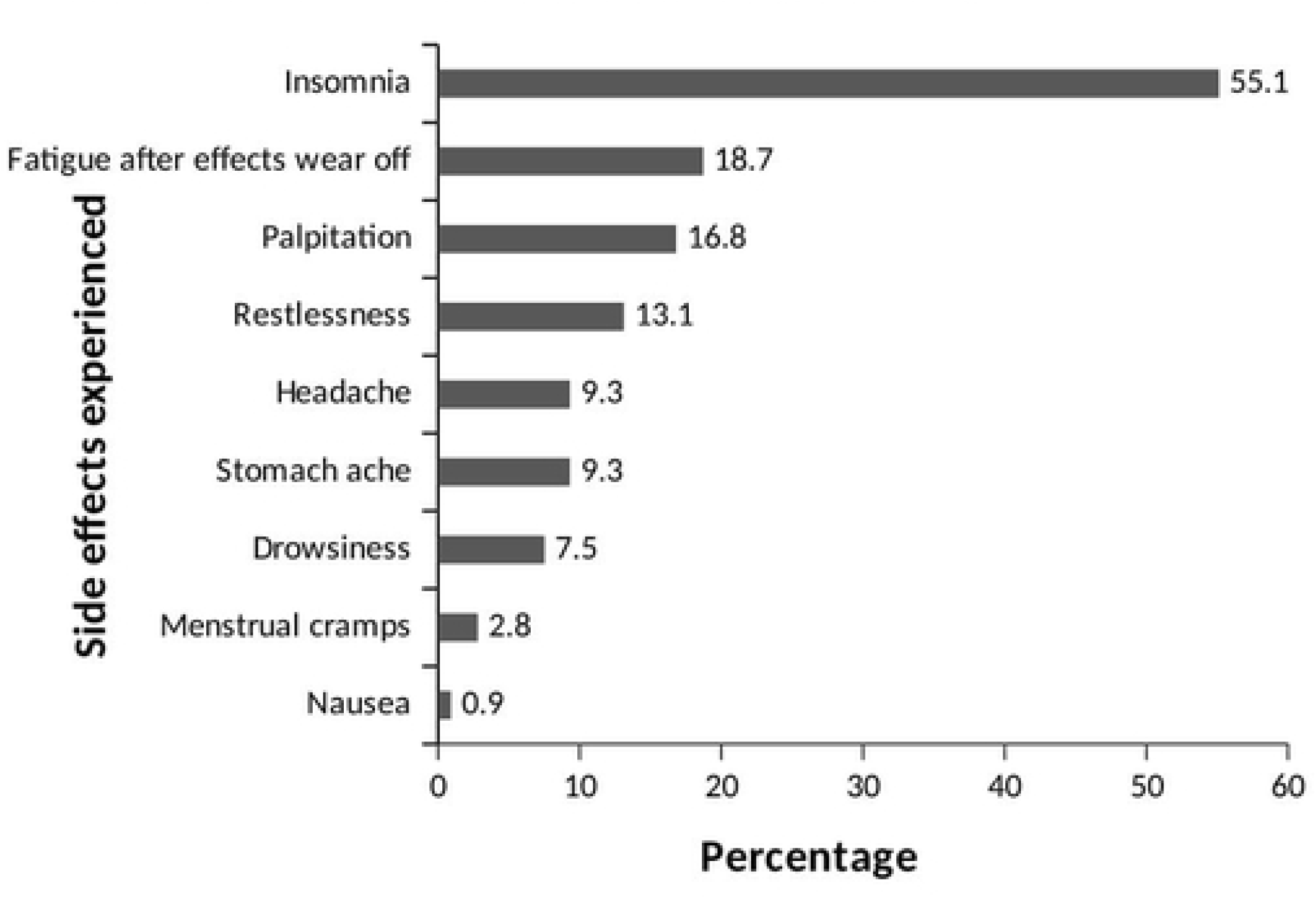
Side Effects Experienced by Energy Drink Users.

**Table 2.** Consumption Pattern Among the Youth in Tamale.

| Variable | Frequency (n) | Percentage (%) |
| --- | --- | --- |
| **Frequency of consumption of energy drink** |  |  |
| Daily | 111 | 26.90 |
| Weekly | 293 | 70.90 |
| Monthly | 9 | 2.20 |
| **Number of bottles drank per sitting** |  |  |
| One | 388 | 93.90 |
| Two | 17 | 4.10 |
| Three | 5 | 1.20 |
| Four | 3 | 0.70 |
| **Duration since consumption commenced** |  |  |
| One year and below | 46 | 11.10 |
| 2 to 3 years | 256 | 62.00 |
| 4 years and above | 11 | 26.90 |
| **Preferred time of consumption** |  |  |
| Morning | 31 | 7.50 |
| Afternoon | 155 | 37.50 |
| Evening/ night | 227 | 55.00 |
| **Reason for preferred time for consumption** |  |  |
| Energy boost for work or studies | 83 | 20.10 |
| To relax or refreshment | 32 | 7.70 |
| No reason | 119 | 28.80 |
| Manage fatigue and stress | 76 | 18.50 |
| Keep awake and concentrate | 56 | 13.60 |
| Craving | 27 | 6.50 |
| For its effects to dissipate by a certain time | 20 | 4.80 |

### Knowledge and perception of energy drink consumption

Almost all of the respondents had heard about energy drinks (97.8%). The source of their information on EDs mainly came from Television (87.3%), friends (83.2%), the internet (51.6%), and radio (41.4%). With regards to known ingredients used for manufacturing EDs, Caffeine (76.4%), Taurine (24.6%), Guarana (24.4%), and sugar (38.4%) were also considerably known compared to lesser possible ingredients such as ginseng (3.6%), and vitamins (5.1%). The popular brand was Rush energy drink (93.2%) (Table 3). The most perceived benefit of taking energy drinks was to provide extra energy (81.0%). A greater proportion (81%) of the respondents perceived that the benefit of energy drinks is to get extra energy and about sixty-two per cent perceive stress reduction as a benefit. About sixty-one per cent of the sample perceived that the adverse effect of energy drinks is insomnia (Table 3). The highest proportion of respondents (83.4%) had poor knowledge of EDs. Those with good and excellent knowledge of EDs were made of comparatively smaller proportions (15.7% and .9% respectively) (Fig 7).

**Fig 7.**
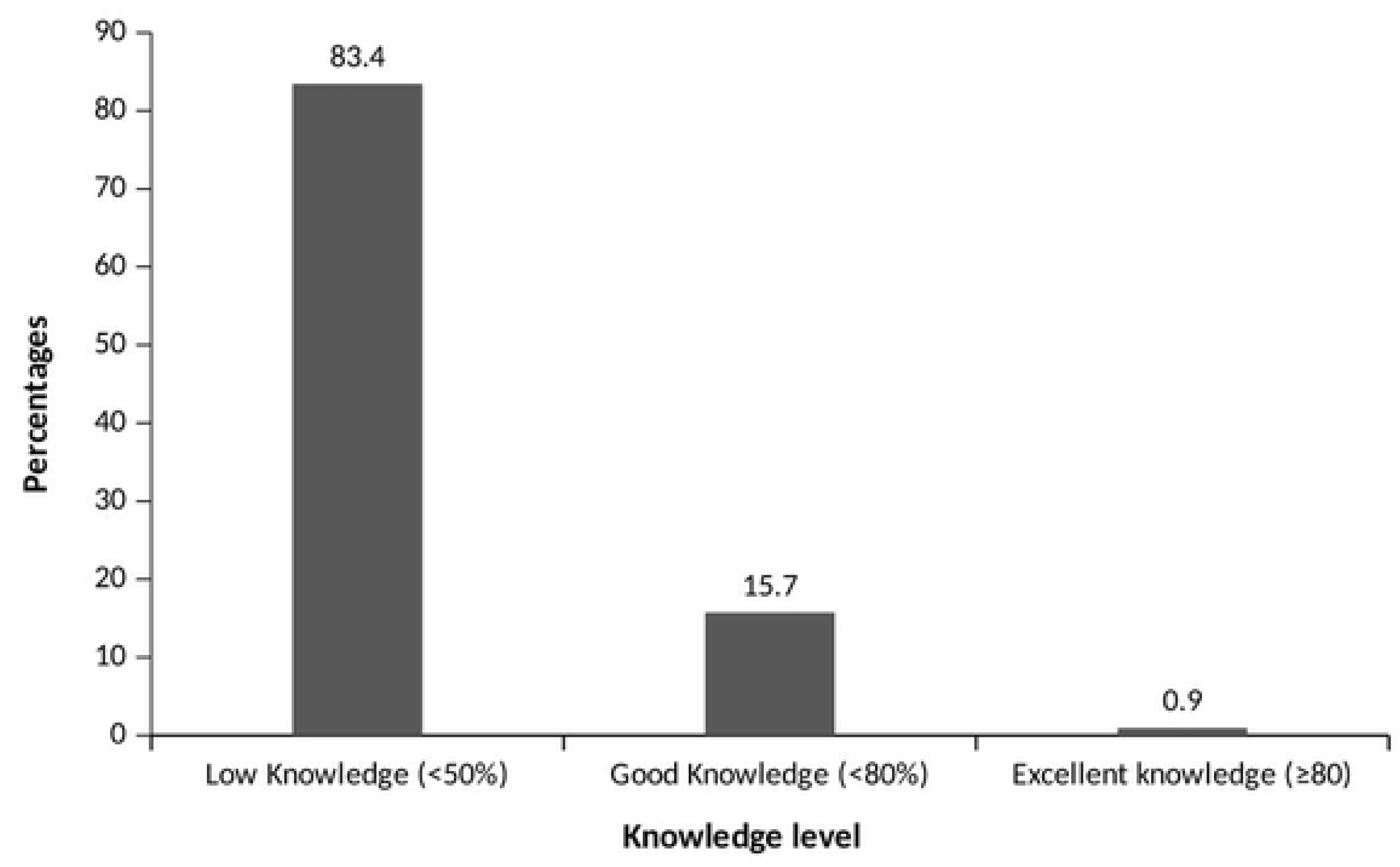
Knowledge Level of Respondents.

**Table 3.** Knowledge and Perception of the Sample on EDConsumption.

| **Variable** | **Frequency (n)** | **Percentage (%)** |
| --- | --- | --- |
| **Ever Heard of Energy Drink** |  |  |
| Yes | 529 | 97.80 |
| No | 12 | 2.20 |
| **Heard About Energy Drinks Through** |  |  |
| Television | 462 | 87.30 |
| Radio | 219 | 41.40 |
| media/internet | 273 | 51.60 |
| Friends | 440 | 83.20 |
| Billboard | 11 | 2.10 |
| **Known Ingredients in Energy Drinks** |  |  |
| Guarana | 129 | 24.40 |
| Taurine | 130 | 24.60 |
| Salt | 24 | 4.50 |
| Caffeine | 404 | 76.40 |
| Water | 326 | 61.60 |
| Sugar | 203 | 38.40 |
| Ginseng | 19 | 3.60 |
| Colour | 6 | 1.10 |
| Flavour | 20 | 3.80 |
| Vitamins | 27 | 5.10 |
| Alcohol | 45 | 8.50 |
| Acid | 19 | 3.50 |
| **Known Brand of Energy Drinks** |  |  |
| Storm | 468 | 88.10 |
| 5-Star | 472 | 88.90 |
| Rush | 495 | 93.20 |
| Burn | 37 | 6.80 |
| Red Bull | 46 | 8.70 |
| Blue Jeans | 119 | 22.40 |
| Tamarinda | 1 | .20 |
| Power Up | 5 | .90 |
| Explode | 10 | 1.90 |
| Rox | 53 | 10.00 |
| Passion | 12 | 2.30 |
| Easy Sport | 123 | 23.20 |
| Run | 146 | 27.30 |
| Lucozade | 31 | 5.80 |
| Vim | 56 | 10.40 |
| 10-10 | 66 | 12.20 |
| Boss | 16 | 3.00 |
| Vody | 26 | 4.90 |
| Bullet | 17 | 3.20 |
| Next Level | 12 | 2.30 |
| **Perceived Benefit of Energy Drink** |  |  |
| Increase work/study concentration | 106 | 20.00 |
| Give extra energy | 430 | 81.00 |
| Boost Appetite | 61 | 11.50 |
| Reduce stress | 331 | 62.30 |
| Nutrient source | 1 | .20 |
| Prevents malaria | 1 | .20 |
| Reduce or manage hunger | 1 | .20 |
| High | 1 | .20 |
| Pain killer | 5 | .90 |
| Sexual stamina | 5 | .90 |
| Aid digestion | 2 | .40 |
| Increase work rate | 1 | .20 |
| Lose weight | 2 | .40 |
| Keep awake | 48 | 9.00 |
| Refreshment | 4 | .80 |
| **Perceived Side Effects of Energy Drinks** |  |  |
| Insomnia | 322 | 60.60 |
| Drowsiness | 59 | 11.10 |
| Restlessness | 273 | 51.40 |
| Headache | 105 | 19.80 |
| Addiction | 20 | 3.80 |

### Relationship between ED use and socio-demographic characteristics

An independent samples T-test revealed that consumers (*M* = 24.4, *SD* = 7.6) were generally younger than non-consumers (*M* = 29.5, *SD* = 8.6), *t*(160.79) = -5.64, *p* < .001. The Chi-square analysis indicated that the proportion of ED consumers decreased across different age categories: 264 (83.8%), 102 (82.9%), and 47 (54%) for the age groups 15-25, 26-35, and 36-45, respectively. Notably, each age group had a higher proportion of consumers compared to non-consumers, and the youngest age category (15-25) exhibited the highest proportion of users.

These frequency differences were statistically significant, *X*^2^(2, N = 525) = 37.8, *p* < .001. The Chi-square test indicated a significantly higher proportion of male consumers (66.6%) compared to female consumers (33.4%), *X*^2^(1, *N* = 525) = 7.36, *p* = .007. Educational level was also found to be significantly associated with consumption, *X*^2^(4, *N* = 525) = 19.14, *p* = .001. Additionally, respondents with high work intensity were more likely to consume energy drinks, *X*^2^(1, *N* = 525) = 24.03, *p* < .001. Although there was no significant association between consumption and alcohol intake, *X*^2^(1, *N* = 525) = 1.299, *p* = .254, a significant association was found between consumption and smoking, *X*^2^(1, *N* = 525) = 6.196, *p* = .013. Non-smokers (96.6%) had a considerably higher proportion of non-consumers, while smokers (3.4%) were more likely to consume energy drinks (Table 4).

**Table 4.**
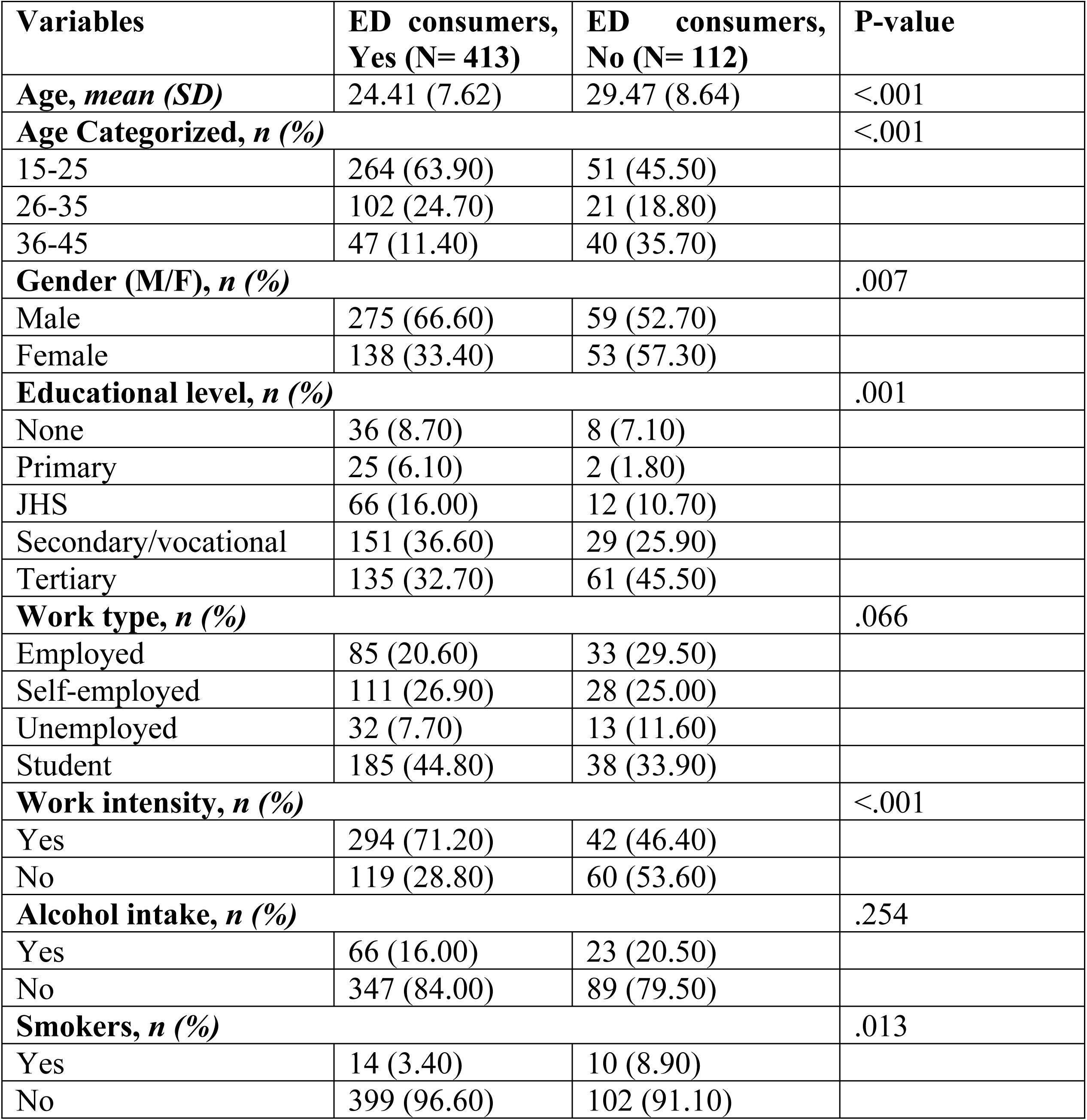
Association Between Energy Drink Use and Socio-Demography.

### Risk factors related to ED consumption

A logistic regression analysis was conducted to examine the predictive value of various factors, including age, gender, marital status, education level, work intensity, alcohol intake, smoking, BMI, energy drink consumption at social gatherings, and knowledge of energy drinks (EDs), on the likelihood of consuming EDs. The logistic regression model yielded significant results, *X*^2^(23, *N* = 525) = 149.17, *p* < .001. The model accounted for 24.7% (Cox & Snell R2) to 38.3% (Nagelkerke R2) of the variance in consumption and accurately classified 83.0% of cases. Table 5 demonstrates that marital status, education level, work intensity, servings at gatherings, and knowledge level of EDs significantly contributed to the model.

**Table 5.** Logistic Regression Predicting the Likelihood of Consuming Energy Drinks.

| Variables |  | OR | 95% C.I. |  | P-value |
| --- | --- | --- | --- | --- | --- |
|  |  |  | Lower | Upper |  |
| Age in categories | 15 – 25 | 1 |  |  | .002 |
|  | 26 – 35 | .359 | .159 | .814 | .014 |
|  | 36 – 45 | 1.515 | .578 | 3.975 | .398 |
| Gender | Male | 1 |  |  |  |
|  | Female | 1.652 | .951 | 2.869 | .075 |
| Marital status | Single | 1 |  |  | .043 |
|  | Married | .872 | .392 | 1.942 | .738 |
|  | Separated | 12.626 | 2.001 | 79.644 | .007 |
|  | Divorced | .925 | .003 | 319.780 | .979 |
|  | Widowed | 3.208 | .354 | 29.027 | .300 |
| Education level | None | 1 |  |  | .001 |
|  | Primary | .465 | .075 | 2.890 | .411 |
|  | JHS | 1.069 | .287 | 3.979 | .921 |
|  | SHS/ vocational | .993 | .316 | 3.124 | .991 |
|  | Tertiary | 3.634 | 1.175 | 11.237 | .025 |
| **Type of work** | Employed | 1 |  |  | .220 |
|  | Self-employed | 1.624 | .747 | 3.532 | .221 |
|  | Unemployed | 1.130 | .398 | 3.208 | .818 |
|  | Student | .700 | .329 | 1.490 | .355 |
| **Work intensity** | Yes | 1 |  |  |  |
|  | No | 3.692 | 2.055 | 6.634 | <.001 |
| **Alcohol intake** | Yes | 1 |  |  |  |
|  | No | .835 | .387 | 1.802 | .646 |
| **Smoking** | Yes | 1 |  |  |  |
|  | No | .672 | .192 | 2.359 | .535 |
| **BMI categories** | Underweight | 1 |  |  | .941 |
|  | Normal | .713 | .242 | 2.098 | .539 |
|  | Overweight | .696 | .206 | 2.356 | .560 |
|  | Obese | .730 | .183 | 2.910 | .655 |
| **EDs served at gatherings** | Yes | 1 |  |  |  |
|  | No | .160 | .092 | .277 | <.001 |
| **Knowledge level** | Low knowledge | 1 |  |  | .010 |
|  | Good knowledge | 2.714 | 1.378 | 5.348 | .004 |
|  | Excellent knowledge | 4.295 | .467 | 39.493 | .198 |

Regarding specific variables, individuals in the 26-35 age group had 0.359 times lower odds of not consuming energy drinks compared to those in the 15-25 age group (*p* = .014, 95% CI .159, .814). On the other hand, individuals in the 36-45 age group had 1.515 times higher odds of not consuming EDs than those in the 15-25 age group (*p* = .398, 95% CI .578, 3.975). Separated individuals had 12.626 times greater odds of not consuming EDs compared to singles (*p* = .007, 95% CI 2.001, 79.644). Furthermore, tertiary-educated individuals had 3.634 times higher odds of not consuming EDs than those without formal education (*p* = .025, 95% CI 1.175, 11.237). Individuals with low work intensity had 3.692 times higher odds of not consuming EDs compared to those with high work intensity (*p* < .001, 95% CI 2.055, 6.634). Moreover, the likelihood of consuming energy drinks was 84% higher when consumed privately compared to being served at public gatherings (*p* < .001, 95% CI .092, .277). Additionally, individuals with good knowledge of EDs were 2.714 times more likely to not consume EDs than those with poor knowledge (*p* = .004, 95% CI 1.375, 5.348). However, age category 36-45, gender, marital status (married, divorced, widowed), work type, alcohol intake, smoking, BMI, and excellent knowledge of EDs did not have a significant impact on the model (Table 5).

## Discussion

Energy drinks are gaining popularity, but studies have linked them to harmful effects. Despite this, more people are consuming them for various reasons. Given the lack of policies in Ghana and aggressive marketing by energy drink companies, it is crucial to understand consumption patterns and predictors. This knowledge can help inform policies and educational efforts by public health professionals to address this growing phenomenon.

According to this research, a high percentage of the study population is aware of energy drinks, consistent with other studies [13,17]. The perceived benefits of these drinks align with their advertised effects, suggesting strong marketing efforts. However, negative effects are often learned through personal experiences rather than advertising. Therefore, obtaining information from friends and relatives can have a more positive impact on decisions about energy drink consumption compared to direct advertising. Other studies have also shown the influence of friends and advertisements on awareness of energy drinks, but the nature of this influence (positive or negative) remains undetermined [13,18].

The high brand awareness among respondents in this study suggests effective advertising and promotion strategies, as well as the visibility of various energy drink brands. Energy drink producers invest heavily in advertising through major outlets such as television, public signs, posters, and sponsorship. Another study found that the popularity of energy drinks is associated with advertising media [19]. Additionally, there is a strong link between energy drink use and exposure to brand advertisements. Advertisements primarily target the youth, who face high demands in their fast-paced lives and may turn to energy drinks as a quick solution. These products are often marketed with strong sports and entertainment themes, leveraging associations with influential celebrities. Advertisements specifically target youthful audiences, with males being the primary focus due to their association with physical activity and sports. Gendered advertising reflects the leveraging of higher involvement of males in sports. Overall, messaging in energy drink advertisements is often associated with sports and entertainment [20].

According to a study by Coso et al. [21], the most common ingredients found in energy drinks, comprising over 50% of the content, are caffeine, vitamin B6, sodium, niacin, and vitamin B12. Caffeine is the most well-known ingredient, with 76.4% awareness among consumers. However, there is a contrast in awareness of other ingredients such as taurine (24.6%), sugar (38.4%), and guarana when compared to similar studies [17]. The popularity of caffeine awareness can be attributed to its prominent marketing as a key ingredient in energy drinks. Lesser-known ingredients are not heavily marketed and may be too technical for consumers to be concerned about. This indicates that consumers of processed products, including energy drinks, prioritize the perceived or real benefits of the product rather than paying attention to its specific contents. Individuals may not fully understand the information provided on energy drink labels or simply place a high level of trust in the producers.

Rush, 5-Star, and Storm are the most popular energy drink brands in the northern region, with high levels of popularity ranging from 88.1% to 93.2%. These findings differ from other studies that identified brands like Black and Red Bull as popular choices. The variation in popularity could be influenced by factors such as economic and demographic conditions, product effectiveness, taste, and availability. The marketing efforts, affordability, and favourable taste of Rush, 5-Star, and Storm brands likely contribute to their popularity in the Tamale Metropolitan area, especially among the youth who prefer these drinks over more expensive premium brands like Red Bull and Blue Jeans.

Energy drinks are perceived to have several benefits, including increased alertness, improved work or study concentration, energy boost, stress management, and promotion of health and social relationships. However, when comparing these perceived benefits to older studies, there are loose similarities. Similarly, the study highlights insomnia, restlessness, headache, and drowsiness as commonly reported adverse effects of energy drinks, aligning with previous research findings [17].

Upon analyzing the data, it is evident that a large proportion of the sample population in this study has poor knowledge of energy drinks (EDs), accounting for 83.4%. This trend is consistent with other studies that have also reported a high rate of poor knowledge among participants [13]. The lack of warning labels and inadequate inclusion of risks and side effects in advertisements and messaging contribute to this poor knowledge [13,22,23]. However, in contrast to previous findings, this study establishes a significant relationship between knowledge level and ED consumption. This suggests that low knowledge is associated with a higher prevalence of ED consumption. It is crucial to address the misconceptions and provide accurate information about EDs, as some respondents even mistakenly associate other carbonated drinks with EDs. Additionally, there is a lack of available information, particularly regarding the adverse effects incorporated in default marketing strategies. Overall, there is a pressing need to improve knowledge and correct misconceptions surrounding EDs among youthful consumers.

### Attitude and practice

Despite the relatively low consumption rate of energy drinks in some environments [24], this study reveals that a significant proportion (78.7%) of the population currently consumes these beverages. This finding aligns with studies conducted in Sicily among adults (78%) and in Ho, Ghana among commercial drivers (75%) [13,17]. Moreover, an even higher percentage of individuals have ever tried energy drinks, with nearly the entire study population (98.7%) reporting consumption at some point. This trend is consistent with studies conducted among adolescents in Shanghai, China (70.5%) and in Ho (85.6%) [13,25,26]. The high prevalence of ever-consuming energy drinks may be attributed to factors such as curiosity and perceived utility. The increasing popularity of energy drinks can be attributed to various factors, including individual factors such as perceived benefits, experienced side effects, and limited knowledge about these beverages. Interpersonal factors like peer pressure, social image, and parental influence also play a role, along with environmental factors such as advertising and branding, affordability, availability, and the lack of policies and regulations.

The main reasons for consuming energy drinks reported by the respondents were performance enhancement, stress management, energy boost, and increased work or study concentration. Similar findings were observed in studies conducted among commercial drivers and students, who relied on energy drinks to manage fatigue, stay awake, and concentrate for extended periods [13,27–29]. Workers using energy drinks to enhance performance may pose risks to themselves and others in the workplace, while students use them to cope with academic pressure. Many energy drink users appreciate their ability to reduce sleep hours, enhance concentration, boost energy, and serve as refreshments. However, non-users tend to avoid them due to awareness of potential side effects [30]. Withdrawal symptoms were reported by energy drink consumers once the effects wore off, potentially leading to dependence and workplace accidents. Some users mix water with energy drinks to reduce their effects, which is seen as a positive health behavior. Adverse effects experienced by consumers included abdominal pain, chest pain, fatigue, constipation, insomnia, palpitations, diuresis, and muscle weakness. Although a wide range of adverse reactions have been reported by energy drink users, this study suggests a small proportion of users experienced side effects, possibly due to responsible consumption patterns within the sample population. Lack of information about adverse health outcomes in advertisements may contribute to negative perceptions based on user experiences, making word-of-mouth information sharing a crucial source of such information for the general public.

Energy drinks are particularly popular among young individuals, especially those in their early twenties, as supported by various studies. This can be attributed to the energy-demanding nature of their activities and the desire to save time. Young people’s curiosity and inclination to follow trends also contribute to their higher consumption of energy drinks compared to older individuals. Additionally, this study found that males generally consumed energy drinks more frequently, particularly at night, and experienced more side effects. These findings align with the marketing strategies of energy drinks, which often target young males and emphasize activities associated with youth and masculinity, such as sports. This focus on young males in energy drink marketing has been observed in multiple studies [20,26,31–34].

The findings of this study demonstrate a clear association between education level and the use of energy drinks (EDs). This relationship may be attributed to the high number of students in the population who rely on EDs for studying purposes. Furthermore, individuals engaged in intense and demanding work activities tend to consume more energy drinks compared to those with less intense work. Intense work is typically characterized by physically demanding tasks, long work hours, and limited breaks. One possible interpretation is that individuals in such occupations find energy drinks to be a convenient way to boost energy without interrupting their work for a meal. This is particularly appealing to young individuals in intense work settings who seek an affordable alternative to food, consume energy drinks quickly, maintain workflow without lengthy breaks, and replenish energy after major meals. It should be noted that while energy drinks may impact energy levels, they may not provide sufficient nutritional value and can lead to decreased reliance on natural foods and inadequate rest management.

On the other hand, individuals with low work intensity are less likely to consume energy drinks since they do not require the same level of energy boost due to their lower energy demands at work. Among energy drink consumers, a small proportion consists of smokers, suggesting that individuals who consume other substances may prefer alternatives to cigarettes. The lack of association between alcohol consumption and energy drink use may be attributed to taste preferences and concerns about the impact of combining the two on performance, which was the primary reason for consuming energy drinks among the participants in this study. Similar to previous studies by Casuccio et al. [17]. and Chang et al. [11], no significant associations were found between ED use and these variables. Furthermore, this study revealed that easy access to energy drinks corresponds to higher consumption rates. Energy drinks are readily available and easily accessible to children and young adults, with numerous brands found in every shop. Additionally, there is a lack of government regulations governing the sale and marketing of energy drinks. This unrestricted promotion and utilization of energy drinks contribute to their increasing appeal, popularity, and availability. The knowledge or experience of side effects also plays a role in influencing the use of energy drinks, as indicated by the study results.

The respondents in this study were primarily within the age range of 15-25 (63.9%), followed by 26-35 (24.7%), and 36-45 (11.4%) age groups. This aligns with previous studies that have shown a lower likelihood of energy drink consumption with increasing age [17,35,36]. It is also consistent with the finding that knowledge about energy drinks tends to increase with age [37].

Among the participants in this study, a high proportion (70.9%) consumed energy drinks on a weekly. A smaller percentage consumed them daily (26.9%) or monthly (2.2%). In contrast, a study by Subaiea et al. [22]. found a higher proportion of monthly consumers and fewer weekly and daily consumers. However, there are differences in the age range and age distribution between the two studies, which may explain the variation in consumption frequency patterns. Other studies have reported that most consumers tend to consume energy drinks weekly and have one drink per day [27,33,38].

Interestingly, the majority of consumers in this study started consuming energy drinks within the past 2 to 3 years. This coincides with the introduction of new and affordable energy drink brands in the Ghanaian market, suggesting that these factors contribute to the increasing popularity of energy drinks. The frequency of consumption is also influenced by the relatively low price of energy drinks, which makes them easily accessible to individuals with lower purchasing power. The data further indicates that the most consumed energy drinks are the cheaper brands such as Storm, Rush, and 5-Star.

While another study suggests that energy drink (ED) consumers do not have a preferred time of consumption, the participants in this study revealed a tendency to consume EDs in the evenings or at night [19]. This finding aligns with the study conducted by Chang et al. [11], where more than half of the consumers reported consuming EDs at night. This preference for evening consumption may be related to specific usage patterns, particularly among those who use EDs to study, stay awake, or manage stress and fatigue after a day of work. Consumers are less likely to consume EDs in the morning when they are already rested and do not require an energy boost. However, consumption rates tend to increase in the afternoon when workers start to feel tired from their morning activities.

ED consumption has been associated with smoking, alcohol intake, cannabis, medical drugs, and opioid use in previous studies [39,40]. However, EDs pose risks to mental health, cardiovascular health, dental health, metabolic health, and social well-being [41,42]. While some studies have reported an association between ED use and alcohol intake [43], this study’s results align with a study by Peacock et al. [31] that found no significant association between ED use and alcohol consumption. The study population in this research consists of few alcohol users and smokers due to the strong religious affiliations among the participants.

Public places and events are the primary locations where EDs are consumed, as reported in previous studies. However, in this study, less than half of the participants mentioned consuming EDs at public gatherings. This differs from another study where a similar proportion of ED users reported consuming them at parties. Special occasions were found to have little influence on the use of EDs according to most users in this study, which contrasts with the findings of Casuccio et al. [17]. When it comes to mixing EDs with other substances, consumption at social gatherings seems to promote such behavior compared to consumption at home [44]. Approximately half of the sightings of EDs at social gatherings in this study were reported at weddings. These results differ from previous literature, possibly due to differences in study populations, demographics, and cultures. It is worth noting that the majority of users and non-users of EDs do not support the idea of mixing them with other substances, as supported by the study conducted by Chang et al. [11]. This is similar to user behavior in this study.

According to the findings of this study, individuals experience various side effects when the effects of energy drinks (EDs) wear off. These side effects include insomnia, fatigue, palpitations, restlessness, headaches, stomach aches, drowsiness, menstrual cramps, and nausea. It is worth noting that these results differ slightly from a previous study conducted by Subaiea et al. [22], which examined the consumption patterns of EDs among the Saudi population. The variation in reported side effects may be attributed to the different brands of energy drinks consumed by each population.

The study explored various factors associated with energy drink (ED) consumption, including staying alert, curiosity, flavor/taste, and concentration on tasks such as school or work [11]. Similar to previous research, this study did not find a significant correlation between ED use and BMI, alcohol intake, or employment status [11,17]. While age was not a significant predictor, there was a decrease in ED consumption with increasing age, consistent with findings that young adults are more likely to consume energy drinks than older adults [45]. Work intensity and education level emerged as predictors of ED use, supporting the notion that activities like work and academics influence calorie intake and, consequently, ED consumption [17,40,46]. Beverage availability was found to be unrelated to consumption [47]. The study’s findings also contradict the claim that gender, smoking, alcohol use, and positive perception of energy drinks predict ED use [11].

### Predictors of energy drink consumption

The findings of this study show that when studying energy drink consumption certain factors that may influence its consumption or otherwise should be looked at in more detail. Age has a relationship with consumption and people in the age category of 15-25 years are less likely to consume EDs compared to those in the 26-35 age category. This may be because the older group of people has purchasing power. They may also tend to consume it more because they feel more drained of energy in their daily lives. This could lead to people turning to EDs to quickly replenish their energy. Meanwhile, younger people may not consume it as much because they have a belief in innate energy levels and may consume it for pleasure and curiosity. On the contrary, in other studies, EDs are consumed among adolescents more than older people [24]. Marital status is associated with energy drink consumption as well. This appears to be the case because relationships with people play a key role in their psychology which encompasses decision-making on nutrition and health-seeking behavior. Therefore, it is not out of place that the findings of this study have revealed marital status to be a factor that influences consumption. The intensity or perceived intensity of work done by an individual inadvertently causes the need for more energy to complete tasks, especially for those in the blue-collar job bracket. This is in line with this study’s findings that indicate that people who perceive their work to be low in intensity are less likely to consume energy drinks. meanwhile, another study conducted among adolescents contradicts this finding by reporting that ED consumption is highest among those who are less physically active [24]. The difference may be a result of a difference in culture in the way adolescents are raised as well as the span of age categories covered by this study in its study population. Education plays a key role in exposure and understanding of information including the types related to health. And this is demonstrated in the findings - the level of education has a relationship with ED consumption. As shown, those with tertiary education are less likely to consume EDs when compared to those who have never had a formal education. This could mean that the exposure to and ability to understand health information such as what EDs are and what their potential side effects may be, people with formal education are better informed to make the right choices. Those who have not had a formal education usually struggle to accept new information and end up accepting myths and discarding truths and facts in some cases. A related factor of interest is the level of knowledge people have of EDs in general. An understanding of what drinks are referred to as EDs, their potential benefits, side effects, and even constituent ingredients could influence the decision to consume ED products. Therefore, special attention should be given to those with low education status and low health literacy because formal and health education could play a critical role in the way in which people consume EDs [33]. Consumption of EDs was found to be 84% higher when consumed in private compared to consumption at public gatherings. This may be the case because public gatherings do not occur often enough for each individual to consume enough EDs. Also being served as a refreshment means that most people will only get a can or a bottle at the said function in addition to the possible availability of other drinks as options for refreshment. On the other hand, it is the private intrinsic needs of individuals that may push them to consume more EDs. Hence, they can easily obtain it more often to satiate that need.

## Conclusions

The study examined the prevalence and consumption of energy drinks (EDs) among the youth in the Tamale Metropolis, along with their perceptions and factors influencing consumption. The findings indicate that more than half of the respondents currently consume EDs. However, there is a lack of knowledge regarding the ingredients, potential benefits, and harmful effects of EDs. Several factors were identified as independently associated with ED consumption, including age, marital status, work intensity, education level, knowledge of EDs, and the availability of EDs at public gatherings. This research highlights the high consumption of EDs and low abuse. This is despite a limited understanding of them, raising concerns about regulations related to advertising, marketing, distribution, and usage, as well as the need to quantify the size of the energy drink market. Unlike previous studies that focused on specific groups such as commercial drivers, students, and athletes, this study provides insights into energy drink use and knowledge within the general population.

## Data Availability

All data produced in the present study are available upon reasonable request to the authors

## Notes

### Competing Interest Statement

The authors have declared no competing interest.

### Funding Statement

This study did not receive any funding.

### Author Declarations

Committee on Human Research, Publications & Ethics (CHRPE) of Kwame Nkrumah University of Science and Technology (KNUST) gave ethical approval for this work

